## Supplementary material for "“*I see salt everywhere*”: A qualitative examination of the utility of arts-based participatory workshops to study noncommunicable diseases in Tanzania and Malawi": S1 Table

| Theme | Description |
| --- | --- |
| Beliefs about NCDs and risk factors | Anything relating to individual perceptions and fears of NCDs incl. • Who is affected? • Urban vs Rural • Perceptions of Dangers and Unpleasantness e.g. death, rotting, scary |
| Impact of NCDs | Anything relating to broader impact of NCDs beyond symptoms • Individual e.g. cannot work • Family e.g. financial instability, stress, dependency • Friends and Neighbours • Government e.g. cost of treatments |
| Experiences of NCD and risk factors | Anything relating to how people have experienced NCDs and risk factors • Personal Accounts • Accounts from Family/Friends/Neighbours etc |
| Causes of NCDs and risk factors | Broad theme including items relating to causes, risk factors and other identified reasons that people acquire NCDS. • diet e.g. too much salt • alcohol consumption • exercise e.g. lack of, laziness • family e.g. inheritance • other e.g. God’s will, financial instability, weakness, chemicals |
| Barriers to NCD Prevention | Anything that could prevent behaviors that could lower risks of NCDs • Food is tasteless without salt/sugar • Financial instability, Costs of healthy food • Exercise: lack of time/planning, lack of safe spaces, can’t be bothered • Social norms incl. perceptions of running • Lack of understanding of NCDs and causes |
| NCD Symptoms | Anything mentioned as an identifying feature of non-communicable diseases, how do NCDs manifest in individuals and how do they vary between different NCDs. e.g. sweating, frequent urination, weakness, wounds |
| *Prevention Facilitators | Counter to Barriers: Anything that currently exist/ are already happening that encourage preventative behaviours |
| *Suggestions for Prevention | Anything to stop people developing NCDs • Diet • Exercise • Raising Awareness/Education: incl. methods e.g. village meetings, arts-based methods, TV etc • Government e.g. improve food regulations, finance farmers to lower prices of health products • Responsibility of food/alcohol vendors e.g. less salt packaging, advising people to drink less |
| *Treatment | Anything relating to how NCDs are treated in current practices including • Biomedicine: hospitals, drugs, costs • Behaviour changes and advise e.g. eat less salt • Traditional: herbal remedies e.g. raw leaf |

*Themes not included in current study
